# Contributions of occupation characteristics and educational attainment to racial/ethnic inequities in COVID-19 mortality

**DOI:** 10.1101/2021.10.29.21265628

**Authors:** Ellicott C. Matthay, Kate A. Duchowny, Alicia R. Riley, Marilyn Thomas, Yea-Hung Chen, Kirsten Bibbins-Domingo, M. Maria Glymour

## Abstract

**Background:** Racial/ethnic inequities in COVID-19 mortality are hypothesized to be driven by education and occupation, but limited empirical evidence has assessed these mechanisms.

**Objective:** To quantify the extent to which educational attainment and occupation explain racial/ethnic inequities in COVID-19 mortality.

**Design:** Observational cohort.

**Setting:** California.

**Participants:** Californians aged 18-65 years.

**Measurements:** We linked all COVID-19-confirmed deaths in California through February 12, 2021 (N=14,783), to population estimates within strata defined by race/ethnicity, sex, age, USA nativity, region of residence, education, and occupation. We characterized occupations using measures related to COVID-19 exposure including essential sector, telework-ability, and wages. Using sex-stratified regressions, we predicted COVID-19 mortality by race/ethnicity if all races/ethnicities had the same education and occupation distribution as White people and if all people held the safest educational/occupational positions.

**Results:** COVID-19 mortality per 100,000 ranged from 15 for White and Asian females to 139 for Latinx males. Accounting for differences in age, nativity, and region, if all races/ethnicities had the education and occupation distribution of Whites, COVID-19 mortality would be reduced for Latinx males (−22%) and females (−23%), and Black males (−1%) and females (−8%), but increased for Asian males (+22%) and females (+23%). Additionally, if all individuals had the COVID-19 mortality associated with the safest educational and occupational position (Bachelor’s degree, non-essential, telework, highest wage quintile), there would have been 57% fewer COVID-19 deaths.

**Conclusion:** Educational and occupational disadvantage are important risk factors for COVID-19 mortality across all racial/ethnic groups, especially Latinx individuals. Eliminating avoidable excess risk associated with low-education, essential, on-site, and low-wage jobs may reduce COVID-19 mortality and inequities, but is unlikely to be sufficient to achieve equity.

## Introduction

As of July 16, 2021, individuals identifying as Latinx were 2·3 times more likely to have died from COVID-19 than non-Latinx White persons in the United States.(1) Inequities for Black and American Indian/Alaska Native groups were similarly large.(1) The drivers of these inequities are poorly understood.(2) Research to date probing the drivers of racial/ethnic inequities in COVID-19 outcomes has focused primarily on risk factors such as age, sex, health status, and comorbidities, all of which inadequately explain racial/ethnic differences in COVID-19 cases, severe morbidity, and mortality.(3–6) A systematic review identified 20 studies examining factors underlying racial/ethnic inequities in COVID-19 outcomes and found they may be partially explained by differences in health care access and factors affecting exposure, but the strength of the evidence was low.(3)

One potential driver of racial/ethnic inequities in COVID-19 outcomes is the unequal distribution of occupations across racial/ethnic groups—one manifestation of structural racism.(7,8) In the US, people of color disproportionately hold jobs that place them at higher risk for poor health outcomes.(8,9) This same pattern has prevailed during the COVID-19 pandemic. Black and Latinx workers were substantially more likely than White and Asian workers to hold low-wage essential jobs and jobs with the greatest risk of COVID-19 exposure relative to population size.(10) In turn, the risk of COVID-19 exposure, infection, and death varies notably by occupation. In California, many non-essential workers experienced no discernable increase in mortality during the pandemic while mortality increased nearly 60% for those in more vulnerable occupations such as sewing machine operators, cooks, and miscellaneous agricultural workers.(11)

Little is known about the contribution of occupation to racial/ethnic disparities in COVID-19 outcomes. One study found that Black and Latinx adults with health conditions that place them at high risk for severe COVID-19 illness were more likely to live in households with essential workers and workers whose jobs cannot be done from home.(12) Several ecological studies suggest that racial/ethnic inequities in COVID-19 outcomes are related to the community-level density of workers in service or low-wage essential occupations, commuting on public transportation, or residing in crowded housing with an essential worker.(13–16) Workplace outbreaks of COVID-19 also appear to disproportionately affect non-White workers.(17) Yet, to our knowledge, no research has directly linked occupation to racial/ethnic inequities in COVID-19 outcomes.

We tested the extent to which racial/ethnic inequities in COVID-19 mortality among the working-age population can be explained by differences in education and occupations. That is, we estimate the magnitude of inequities that would remain if all racial/ethnic groups had similar education and were equally likely to hold low-risk and high-risk jobs. We focus on California, a state of 40 million residents, the most COVID-19 deaths of any USA state, and large racial/ethnic inequities in COVID-19 mortality.(18) Because structural racism manifests not only in the jobs individuals hold, but also in the educational opportunities that determine job opportunities, we also consider the role of educational attainment in racial/ethnic inequities in COVID-19 outcomes. Because both the types of work available and the risk associated with work differ for males and females within racial/ethnic groups, we stratify all analyses by sex.(19,20)

## Methods

### Death data and measures

From California death records, we identified all COVID-19-confirmed deaths occurring between January 1, 2020 and February 12, 2021. Records included the decedent’s race/ethnicity, sex (male or female), date of birth, date of death, and open text fields for primary occupation and industry, described as “type of work done during most of working life”. COVID-19 deaths were those with a primary International Classification of Diseases, Revision 10 diagnostic code of U071.

We conceptualized race/ethnicity as socially defined categories that govern the distribution of risk, opportunities, and discrimination according to each group’s socially-assigned value and power.(21) We classified race/ethnicity as: Asian, Black, Latinx, White, and Other (including American Indian, Alaskan Native, Native Hawaiians, other Pacific Islanders, multi-race, and unspecified). Apart from Latinx, all racial/ethnic groups were non-Latinx. We used the National Institute for Occupational Safety and Health’s Industry and Occupation Computerized Coding System, an automated machine learning-based system, to convert the open text fields for occupation and industry to standardized 2010 Census Codes. Educational attainment categories were: no high school degree and no GED; high school degree or GED; some college or Associate’s degree; and Bachelor’s degree or beyond. Nativity was USA-born or foreign-born. Place of residence was grouped into ten California regions (Appendix Table 1). Given our focus on workers, we restricted to decedents aged 18-65 years at death, in alignment with prior research.(11) We calculated age using dates of birth and death and defined age groups: 18-24 years, 5-year age groups between ages 25 and 59 years, and 60-65 years.

We defined population strata by cross-classifying all categories of all variables selected from the death records. The cross-classification of race/ethnicity, sex, age group, nativity, region, education, and occupational category (see below) created 3,672,000 total possible strata, of which 12,850 were represented in the death data. We then created a dataset comprised of stratum-level COVID-19 death counts by summing the number of COVID-19 deaths in each stratum.

### Population data and measures

To characterize the population at risk of death, we used the 2019 American Community Survey (ACS) California person-level microdata. ACS is the primary national population surveillance system, surveying 3 million individuals annually, including 380,000 in California. Residents are legally required to respond. We defined strata using the same set of variables as in the death records. We converted 2018 Census occupation codes in the ACS to 2010 Census occupation codes using the Census Bureau crosswalk. Restricting to the same ages (18-65 years), we created population counts by summing the ACS person weights representing the number of people in each stratum (174,315 strata had non-zero populations).

### Occupational sector and characteristics

We characterized occupations using multiple measures hypothesized to be related to SARS-CoV-2 exposure risk (Appendix “Occupational measures”; Appendix Table 2). First, as in previous research,(11) a team of three researchers manually categorized the 529 unique 2010 Census occupation codes into 9 occupational sectors based on the California official definition of essential work(22): facilities, food/agriculture, government/community, health/emergency, manufacturing, retail, transportation/logistics, not essential, and unemployed/not in labor force/missing.

Second, we used Dingel and Neiman’s classification of which jobs can be done at home during the COVID-19 pandemic to link occupation codes to their telework-ability. This classification was based on a composite of job characteristics measures in the Bureau of Labor Statistics O*NET database. In secondary analyses, we also considered 13 individual O*NET measures (Appendix “Occupational measures”).

Third, we linked each occupation code to its median annual wages as reported by the Bureau of Labor Statistics in May 2019. Individuals with lower incomes have less ability to forgo work or income when faced with undesired COVID-19 exposure risk. In secondary analyses, we considered other quantiles of annual and hourly wages for each occupation code. To merge the telework, O*NET, and wages measures to the death/population data, we used multiple occupation code crosswalks (Appendix “Occupation code crosswalks”).

### Construction of analytic dataset

We treated the state of California as a cohort and appended (i.e. stacked) the death and ACS population data to create one strata-level dataset representative of the cohort experience. The outcome was COVID-19 death: rows derived from the death data were assigned a value of 1 and rows derived from the ACS data were assigned a value of 0. Each row was assigned a weight corresponding to the number of individuals it represented, i.e., within a given stratum, the number of individuals who died or who lived in California. These weights were included in all statistical analyses. Because the ACS does not indicate which individuals subsequently died of COVID-19, this data structure implies that the <0·1% of Californians aged 18-65 who died of COVID-19 are represented twice. Records with missing education or occupational measures were excluded (<1% of study population). The final analytic dataset included 187,165 rows representing 25,235,092 individuals aged 18-65 years.

### Statistical analysis

Consistent with prior research studying sources of racial/ethnic health inequities, we treat race/ethnicity as the exposure, educational and occupational variables as potential mediators, and COVID-19 death as the outcome.(23) We conducted sex-stratified analyses using g-computation.(24) This approach allowed us to predict COVID-19 mortality for hypothetical alternative distributions of covariates and educational/occupational positions while flexibly modeling COVID-19 mortality risk.

We fit three separate linear probability models:(25) First, we fit unadjusted linear regression models, regressing COVID-19 death on race/ethnicity. Second, we adjusted for measured compositional differences between racial/ethnic groups (age group, nativity, region). Third, we further adjusted for the hypothesized mediating variable(s) (education, essential sector, telework, wages, or all four measures simultaneously). To allow for non-additive and non-linear associations, we converted all continuous measures to quintile-based categorical measures and included all possible first-order interaction terms between independent variables. For occupational mediators, nonworkers (16% of the study population) were included as a distinct category.

Using the fitted models, we predicted the COVID-19 mortality risk for each racial/ethnic group under four scenarios:

1. Unadjusted: the observed COVID-19 mortality risk predicted using the unadjusted models
2. Composition-adjusted: the COVID-19 mortality risk if all racial/ethnic groups had the same distribution of covariates (age, nativity, and region) as Whites (predicted using the covariate-adjusted models)
3. Composition- and mediator-adjusted: the COVID-19 mortality risk if all racial/ethnic groups had the same distribution of covariates and education/occupation mediator(s) as Whites (predicted using the covariate- and mediator-adjusted models)
4. Composition-adjusted with safest education/occupation: the COVID-19 mortality risk if all racial/ethnic groups had the same distribution of covariates as Whites and if all individuals, regardless of race/ethnicity, had been in the lowest-risk categories of education/occupation (i.e. Bachelor’s degree or higher, non-essential, telework-able, highest quintile of median annual wages) (predicted using the covariate- and mediator-adjusted models).

This sequence of estimates quantifies how COVID-19 mortality would change for each racial/ethnic group under each hypothetical scenario. To convert to potential deaths averted, we multiplied differences in risk by corresponding population counts. We do not report confidence intervals because the study population constitutes the entire population of interest, not a sample from the population to which we made inferences. Because linear probability models can predict risks outside of [0,1], we tested the sensitivity of our results to using logistic regression.

### IRB approval

This study was approved by the institutional review boards of the California Department of Public Health and the University of California, San Francisco.

## Results

Among people in California aged 18-65 years, there were 14,783 deaths attributed to COVID-19 between January 1, 2020 and February 12, 2021. With an estimated 25 million people at risk, this implies a COVID mortality risk of 59 per 100,000 persons. By design, the demographic composition of the study population matched that of the 2019 California population (Table 1). Compared to the general population, COVID-19 decedents were disproportionately older, male, and Latinx, with lower education and occupational positions (Table 1). For example, those with a high school education or less made up 36% of the study population but comprised 69% of COVID-19 deaths, while those holding jobs that are likely to be telework-able also made up 36% of the population but comprised only 18% of COVID-19 deaths.

**Table 1:** Demographic, educational, and occupational characteristics of 2019 California population and California COVID-19 decedents ages 18-65 years, January 1, 2020-February 12, 2021

| Characteristic | California Population |  | California COVID-19 deaths |  |
| --- | --- | --- | --- | --- |
|  | *N* | % | *N* | % |
| Persons | 25,235,092 | 100% | 14,783 | 100% |
| Age (years) |  |  |  |  |
| 18-24 | 3,688,420 | 15% | 82 | 1% |
| 25-44 | 11,347,306 | 45% | 1,852 | 13% |
| 45-65 | 10,199,366 | 40% | 12,849 | 87% |
| Sex |  |  |  |  |
| Female | 12,514,931 | 50% | 4,549 | 31% |
| Male | 12,720,161 | 50% | 10,234 | 69% |
| Race/ethnicity |  |  |  |  |
| Asian | 3,925,494 | 16% | 1,118 | 8% |
| Black | 1,472,151 | 6% | 966 | 7% |
| Latinx | 9,859,259 | 39% | 10,229 | 69% |
| Other | 937,809 | 4% | 472 | 3% |
| White | 9,040,379 | 36% | 1,998 | 14% |
| Foreign-born |  |  |  |  |
| Yes | 8,305,063 | 33% | 8,808 | 60% |
| No | 16,929,792 | 67% | 5,738 | 39% |
| Missing | 237 | 0% | 237 | 2% |
| Educational attainment |  |  |  |  |
| No HS degree and no GED | 3,633,198 | 14% | 5,377 | 36% |
| HS degree or GED | 5,594,804 | 22% | 4,814 | 33% |
| Some college or Assoc. degree | 7,882,332 | 31% | 2,595 | 18% |
| Bachelor's degree or higher | 8,124,163 | 32% | 1,402 | 9% |
| Missing | 595 | 0% | 595 | 4% |
| Worker sector |  |  |  |  |
| Facilities | 2,561,383 | 10% | 2,715 | 18% |
| Food and agriculture | 1,888,544 | 7% | 1,504 | 10% |
| Government & community | 2,313,800 | 9% | 920 | 6% |
| Health or emergency | 2,025,529 | 8% | 1,006 | 7% |
| Manufacturing | 1,157,573 | 5% | 1,394 | 9% |
| Retail | 1,583,633 | 6% | 698 | 5% |
| Transportation & logistics | 1,805,127 | 7% | 2,003 | 14% |
| Not essential | 7,805,001 | 31% | 2,029 | 14% |
| Unemployed/not in labor force | 4,094,502 | 16% | 1,745 | 12% |
| Missing | 0 | 0% | 769 | 5% |
| Telework-able occupation |  |  |  |  |
| Yes | 8,957,808 | 35% | 2,681 | 18% |
| No | 11,773,666 | 47% | 9,351 | 63% |
| Unemployed/not in labor force | 4,094,495 | 16% | 2,507 | 17% |
| Missing | 409,123 | 2% | 244 | 2% |
| Median annual wage for occupation |  |  |  |  |
| \$22,200 – 29,000 | 4,722,209 | 19% | 3,003 | 20% |
| \$29,001 – 39,100 | 4,130,764 | 16% | 3,405 | 23% |
| \$39,101 – 51,700 | 3,949,646 | 16% | 3,068 | 21% |
| \$51,701 – 73,800 | 3,969,152 | 16% | 1,677 | 11% |
| \$73,800 + | 4,196,965 | 17% | 1,068 | 7% |
| Unemployed/not in labor force | 4,094,495 | 16% | 2,507 | 17% |
| Missing | 171,861 | 1% | 55 | 0% |

Abbreviations: HS: High school. Assoc.: Associate's.

Table 2 presents unadjusted COVID-19 mortality by race/ethnicity and sex, and predicted mortality if all groups had the same composition (age, nativity, region of residence), education, or occupational risk as White people of the same sex (for risk differences, see Appendix Table 3). In unadjusted analyses, Latinx, Black, and other race/ethnicity groups had greater COVID-19 mortality than White people of the same sex. Gaps were particularly pronounced for Latinx people. For Asian females, COVID-19 mortality matched that of White females. Large differences in mortality persisted even after accounting for compositional differences in age, nativity, and region of residence.

**Table 2:** Predicted COVID-19 mortality risks and percent change in mortality risk after accounting for racial/ethnic differences in composition, education, and occupational mediators, for individuals aged 18-65, per 100,000 persons, by race/ethnicity and sex, California, January 1, 2020-February 12, 2021

| Sex,<br>race/<br>ethnicity | Unadjusted<br>COVID-19<br>mortality<br>risk | Composition-adjusted<br>COVID-19<br>mortality risk:<br>if all<br>racial/ethnic<br>groups had<br>the same<br>composition<br>as Whites | COVID-19<br>mortality risk if<br>all racial/ethnic<br>groups had the<br>same<br>composition and<br>education<br>distribution as<br>Whites<br>(% change<br>relative to<br>composition-adjusted) | COVID-19<br>mortality risk if<br>all racial/ethnic<br>groups had the<br>same<br>composition and<br>work sector as<br>Whites<br>(% change<br>relative to<br>composition-adjusted) | COVID-19<br>mortality risk if<br>all racial/ethnic<br>groups had the<br>same<br>composition and<br>telework capacity<br>as Whites<br>(% change<br>relative to<br>composition-adjusted) | COVID-19<br>mortality risk if<br>all racial/ethnic<br>groups had the<br>same<br>composition and<br>wages<br>distribution as<br>Whites<br>(% change<br>relative to<br>composition-adjusted) | COVID-19<br>mortality risk if<br>all racial/ethnic<br>groups had the<br>same<br>composition,<br>education, work<br>sector, telework<br>capacity, and<br>wages as Whites<br>(% change<br>relative to<br>composition-adjusted) |
| --- | --- | --- | --- | --- | --- | --- | --- |
| Female |  |  |  |  |  |  |  |
| White | 15 | 15 | 15 (0%) | 15 (0%) | 15 (0%) | 15 (0%) | 15 (0%) |
| Latinx | 60 | 70 | 55 (-21%) | 69 (-1%) | 68 (-3%) | 66 (-6%) | 54 (-23%) |
| Black | 48 | 51 | 47 (-8%) | 52 (+2%) | 52 (+2%) | 53 (+4%) | 47 (-8%) |
| Asian | 15 | 13 | 17 (+31%) | 13 (0%) | 13 (0%) | 14 (+8%) | 16 (+23%) |
| Other | 29 | 46 | 41 (-11%) | 44 (-4%) | 45 (-2%) | 43 (-7%) | 42 (-9%) |
| Male |  |  |  |  |  |  |  |
| White | 26 | 26 | 26 (0%) | 26 (0%) | 26 (0%) | 26 (0%) | 26 (0%) |
| Latinx | 138 | 141 | 116 (-18%) | 127 (-10%) | 132 (-6%) | 128 (-9%) | 110 (-22%) |
| Black | 77 | 87 | 77 (-11%) | 96 (+10%) | 96 (+10%) | 88 (+1%) | 86 (-1%) |
| Asian | 41 | 27 | 34 (+26%) | 27 (0%) | 26 (-4%) | 27 (0%) | 33 (+22%) |
| Other | 57 | 83 | 77 (-7%) | 81 (-2%) | 81 (-2%) | 80 (-4%) | 80 (-4%) |

Legend: CI: confidence interval. Composition-adjusted rates indicate the predicted COVID-19 mortality risk if all racial/ethnic groups had the same distribution of age, nativity, and region of residence as Whites.

If all groups had the same composition and educational attainment as White people of the same sex, we predicted that COVID-19 mortality would be reduced (and therefore more equal) for Latinx females (−21%) and males (−18%), Black females (−8%) and males (−11%), and Other race/ethnicity females (−11%) and males (−7%). However, COVID-19 mortality inequalities would be exacerbated for Asian females (+31%) and males (+26%), reflecting that on average Asian people in California are more educated than Whites (Appendix Table 4). If all groups had the same composition and work sector or telework capacity as Whites, we predicted little change in COVID-19 mortality for females, but reduced mortality for Latinx males (6-10%) and increased mortality for Black males (+10%). The latter finding is consistent with the pattern that although Black people aged 18-65 in California on average hold far fewer non-essential jobs than Whites (26% vs. 40%), they are also more likely to be unemployed or not in the labor force (23% vs. 14%) (Appendix Table 4) which appears protective against COVID-19 mortality (Table 1).

When equalizing wages, we predicted reduced COVID-19 mortality for Latinx females and males and other race/ethnicity females, but greater mortality for Asian males and little change for other groups. Models adjusting for other occupational measures showed attenuated but similar patterns (Appendix Table 5).

Taken together, if each group had the education, essential sector, telework, and wages of their White counterparts, COVID-19 mortality would be reduced by 23% for Latinx females, 22% for Latinx males, and smaller amounts for Black and other/race ethnicity individuals, but increased by 23% for Asian females and 22% for Asian males (Table 2). Findings based on logistic regression rather than linear probability models were consistent with the main results (Appendix Table 6).

Figure 1 summarizes the unadjusted and predicted COVID-19 mortality risk by race/ethnicity and sex under alternative distributions education and occupation. If all groups held the safest educational and occupational positions (Bachelor’s degree or higher, non-essential, telework-able occupation, highest quintile of median annual wages), we predict that COVID-19 mortality risk would be substantially reduced for all groups, with reductions ranging from 3 per 100,000 among Asian and White females to 75 per 100,000 among Latinx males.

**Figure 1:**
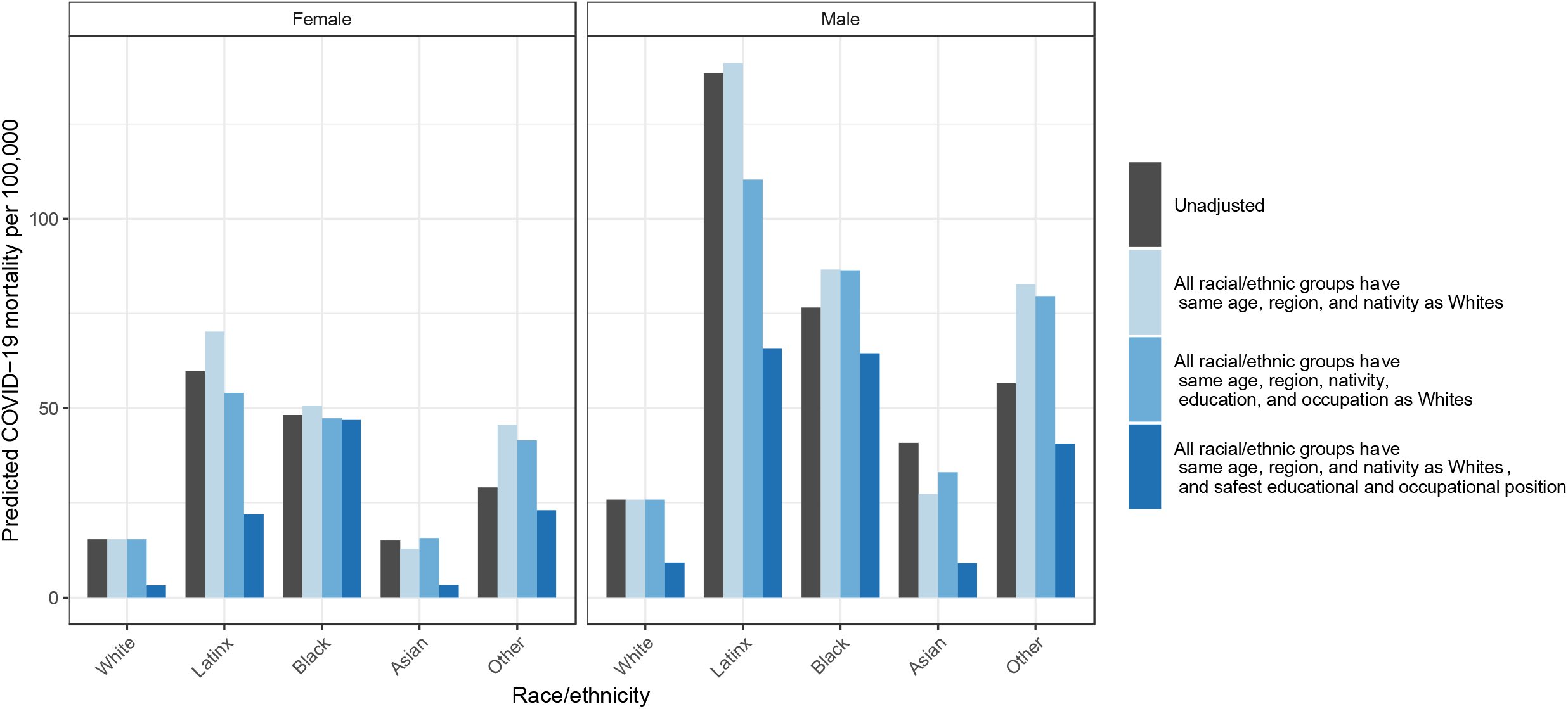
Unadjusted and predicted COVID-19 mortality for individuals aged 18-65, per 100,000 persons, by race/ethnicity and sex, under alternative compositional, educational, and occupational distributions, California, January 1, 2020-February 12, 2021 Legend: Estimates present the predicted COVID-19 mortality risk for each racial/ethnic-sex group under four scenarios: Unadjusted (the COVID-19 mortality risk as-observed); if all racial/ethnic groups had the same distribution of age, nativity, and region of residence as Whites; if all racial/ethnic groups had the same distribution of age, nativity, and region of residence, education, essential sector work, telework-able occupation, and wages as Whites; and if all racial/ethnic groups had the same distribution of age, nativity, and region of residence as Whites and if all individuals regardless of race/ethnicity had a Bachelor’s degree or higher and worked in non-essential, telework-able occupations in the highest quintile of median annual wages.

Table 3 presents predicted deaths among working-age Californians averted by placing everyone in the safest educational and occupational positions. If every working-age Californian had had the COVID-19 mortality risk associated with the safest education and occupational strata, mortality would have decreased substantially for all groups, with the largest relative benefits for White females (−80%) and White males (−65%). The smallest relative benefits would have accrued to Black women, for whom mortality would have been only 8% lower. Overall, this shift was associated with 8,441 COVID-19 deaths averted, a 57% reduction, with the vast majority of this risk reduction among Latinx males (3,755 potential deaths averted) and females (2,329 potential deaths averted).

**Table 3:**
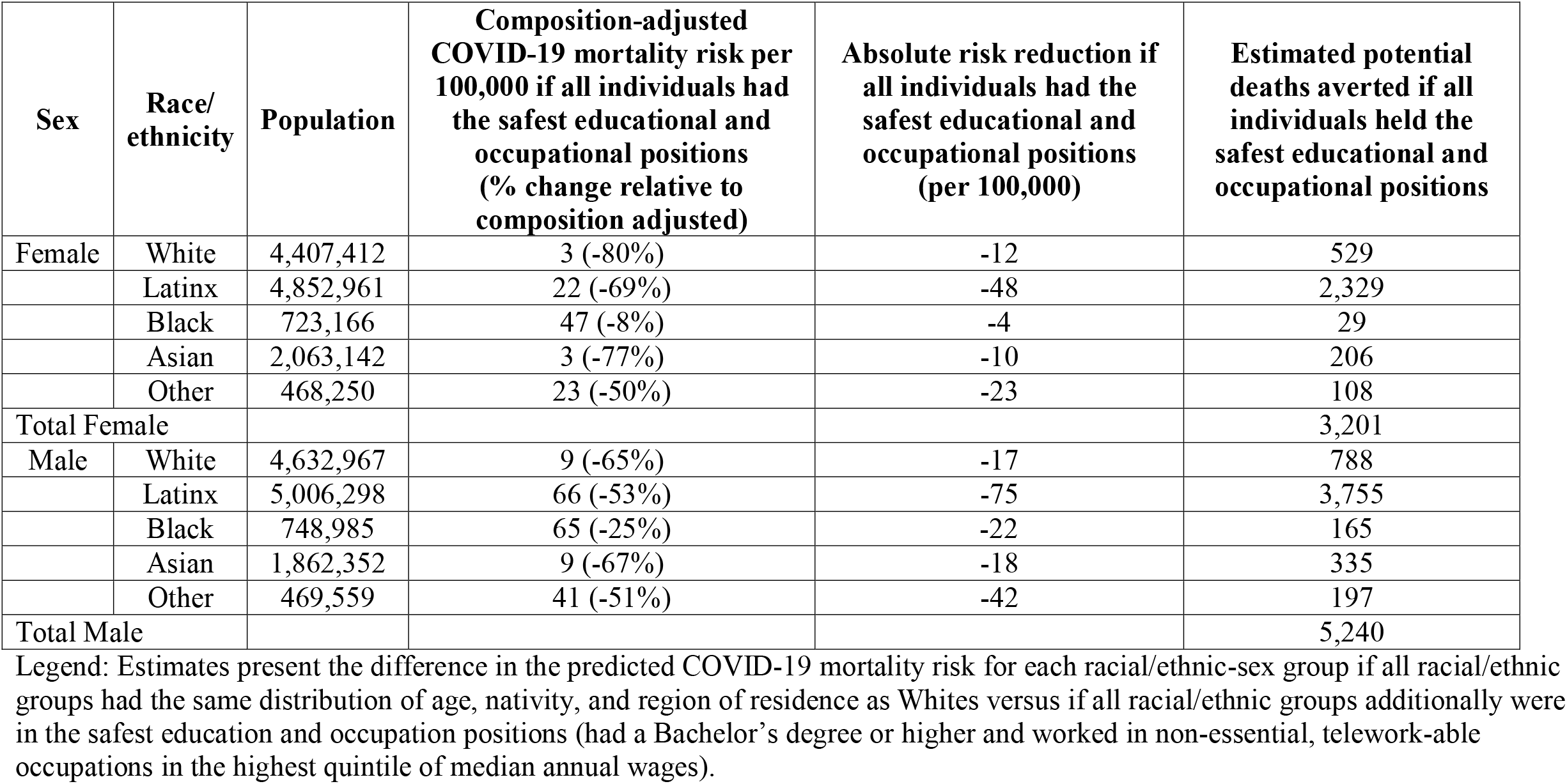
Predicted COVID-19 mortality reductions and potential deaths averted, for individuals aged 18-65, per 100,000 persons, by race/ethnicity and sex, if all groups held the safest educational and occupational positions, California, January 1, 2020-February 12, 2021

## Discussion

In this population-based analysis of working-age Californians, we found that large inequities in COVID-19 mortality by race/ethnicity can be partially explained by differences in educational attainment and measured occupational risk factors. The fraction of inequality in comparison to White people that was explained by education and occupation varied by race/ethnicity and sex. For Latinx males, 22% of their excess risk compared to White males might have been eliminated if they shared the risk associated with the education and occupational conditions of White males; for Latinx females, this number was 23%. For Asian individuals, existing educational and occupational differences apparently conferred an advantage compared to Whites. In all racial/ethnic groups, COVID-19 mortality would have been reduced substantially reduced if excess risk associated with educational and occupational disadvantage were eliminated. If COVID-19 mortality were no higher in low-education, essential, on-site, or low-wage jobs than in high-education, non-essential, telework-able, high-paying jobs, we estimate that around 57% of California’s working-age COVID-19 deaths would have been prevented. In terms of absolute deaths averted, this risk reduction would confer the most benefit for Latinx males and females. Despite these reductions, substantial racial/ethnic inequities in COVID-19 mortality remained, suggesting that eliminating excess risk associated with educational and occupational disadvantage alone is insufficient, and should be considered in complement to other steps to reduce inequities.

Although these findings suggest that education and occupation play important roles in COVID-19 risk and inequities, the estimated percentage of inequity explained was smaller than we anticipated. Compared to measured occupational characteristics, education played a larger role in racial/ethnic inequities in COVID-19 mortality. Prior work has shown that lower educational attainment is associated with greater COVID-19 infection and mortality.(26–28) Inequities in education are driven by residential segregation, educational access, structural racism,(29) and immigration patterns. Education may influence COVID-19 mortality inequities because lower-educated individuals have less access to accurate information on COVID-19 prevention compared to people with more education.(30) Additionally, in the USA, education is tied to employment opportunities, which influence access to healthcare.(31) Healthcare access has been linked to racial/ethnic disparities in COVID-19 outcomes,(3) and despite reductions in racial/ethnic gaps in healthcare coverage after the Affordable Care Act, Black and Latinx adults remain less likely to be covered.(32) Education also conveys social status, social influence, and labor market resources. As a result, individuals with more education may be able to negotiate for safer environments at work or elsewhere, and refuse to participate in unsafe workplaces.(33)

Our findings also underscore the importance of safeguarding essential workers in the highest-risk, lowest-wage occupations. The unequal distribution of risky work contributed to racial/ethnic mortality differences, particularly affecting Latinx individuals. We know how to protect workers: in California, those with high exposure risk but adequate protective policies, procedures, and equipment (e.g. physicians) did not experience increases in pandemic-era mortality.(11) This is especially important as novel pathogenic variants continue to evolve and many remain unvaccinated: despite highly efficacious vaccine availability, as of June 30, 2021, Latinx Californians constituted 63% of COVID-19 cases but only 29% of vaccinations received.(34)

The particular relevance of occupational factors (e.g., essential work, telework, wages) for Latinx people may reflect occupational segregation in California, where the diverse Latinx population is highly differentiated by type of work and corresponding COVID-19 risk. Prior research has shown that Latinx people, in particular, are overrepresented in low-wage, precarious work in the essential sectors that saw the highest COVID-19 mortality in the state.(35) In contrast, our occupational measures did little to explain Black-White inequities. Given the pervasiveness of anti-Black racism, other social factors such as residential segregation, lack of healthcare access, intergenerational wealth inequalities, and less-measurable influences may contribute more to underlying risk for Black workers.

Our findings underscore the importance of considering structural drivers of racial/ethnic inequities in COVID-19 outcomes. A major barrier to research on the role of upstream social and economic factors is the lack of surveillance data that incorporates information on race/ethnicity, education/occupation, and COVID-19 outcomes simultaneously. California death certificates are one of few sources with complete data on all three. Adequate surveillance data on modifiable determinants of inequities such as occupational exposures is urgently needed to advance research on COVID-19 disparities and guide public health efforts to prevent unnecessary COVID-19 deaths.

This study has several limitations. First, we may underestimate the relevance of education and occupation because we used indirect measures of risk that were indexed by codes for primary occupation in life rather than direct measures of occupational exposure. Second, we did not account for within-household transmission initiated by an occupational exposure. We may underestimate the importance of occupation since household members with different occupation codes share COVID-19 risk associated with each other’s work. Third, we only included confirmed COVID-19 deaths, conceivably leading to an underestimate of true inequities: some racial/ethnic groups may be more likely to die at home without COVID-19 testing and therefore not be counted as a COVID-19-confirmed death.(36) Fourth, we could only control for potential confounders measured in both the death and ACS records. Absent further covariate adjustment, education and occupation may proxy for other factors such as social class, intergenerational wealth/debt, parental education, and non-citizen legal status.(37) Unmeasured potential confounders of particular concern are comorbidities, housing composition and density (including housing instability, homelessness, and incarceration), access to high-quality healthcare, and undocumented legal status. However, these factors may also be part of the causal pathway from structural racism, education, and occupation to COVID-19 death.

## Conclusions

Educational attainment and occupation should be considered important risk factors for COVID-19 mortality. Even in one of America’s most progressive states, many people of color have been relegated to high-risk, devalued, unprotected jobs characterized by inadequate COVID-19 protections and limited resources and power to advocate for better protections. Unnecessary COVID-19 mortality is just one of many potential health and social consequences of this racial stratification.(7,8) Future COVID-19 mitigation strategies should include policies, protections, and vigilant monitoring such that workers in low-education, essential, on-site, and low-wage jobs are ensured no greater risk of COVID-19 mortality than workers with greater power and privilege. These steps are not a panacea, but they have the potential to save lives and reduce some racial/ethnic inequities in COVID-19 mortality.

## Data Availability

Death data used for this analysis include person identifiers and cannot be shared publicly. They can be obtained directly from the California Department of Public Health with appropriate approvals. American Community Survey microdata are publicly available online from https://data.census.gov.

## Appendix

### Supplemental Methods

**Appendix Table 1:** Definition of California regions.

| Region number | Counties included |
| --- | --- |
| 1 | Butte, Colusa, Del Norte, El Dorado, Glenn, Lassen, Modoc, Nevada, Placer, Plumas, Sacramento, Shasta, Sierra, Siskiyou, Sutter, Tehama, Trinity, Yolo, Yuba |
| 2 | Humboldt, Lake, Mendocino, Napa, Sonoma |
| 3 | Alameda, Contra Costa, Marin, San Francisco, San Mateo, Santa Clara, Solano |
| 4 | Alpine, Amador, Calaveras, Madera, Mariposa, Merced, Mono, San Joaquin, Stanislaus, Tuolumne, Inyo |
| 5 | Monterey, San Benito, San Luis Obispo, Santa Barbara, Santa Cruz, Ventura |
| 6 | Fresno, Kern, Kings, Tulare |
| 7 | Riverside, San Bernardino |
| 8 | Los Angeles |
| 9 | Orange |
| 10 | Imperial, San Diego |

#### Occupational measures

We characterized occupations using multiple measures hypothesized to be related to SARS-CoV-2 exposure risk (Appendix Table 1). First, as in previous research,^1^ a team of three researchers manually categorized the 529 unique 2010 Census occupation codes into 9 occupational sectors based on the California official definition of essential work^2^ and retail work. The categories were: facilities, food/agriculture, government/community, health/emergency, manufacturing, retail, transportation/logistics, not essential, and unemployed/not in labor force/missing.

Second, we used the O*NET database to link occupation codes to characteristics of each occupation. Overseen by the Bureau of Labor statistics, O*NET is based on surveys completed by employees, employers, and job experts, and includes measures of required knowledge and skills, typical tasks, exposures encountered, and the workplace environment for nearly 1,000 occupations. We used 13 O*NET measures deemed relevant to COVID-19 exposure risk in previous research^3–6^—for example, the importance of assisting and caring for others, or the importance of working with computers (Appendix Table 2). We also considered Dingel and Neiman’s classification of which jobs can be done at home during the COVID-19 pandemic (telework), which was based on a composite of O*NET measures.^5^

Third, individuals with lower incomes have less ability to not work or forgo income when faced with undesired COVID-19 exposure risk. We therefore characterized mean, median, 10^th^, 25^th^, 75^th^, and 90^th^ percentiles of annual and hourly wages within each occupation code, based on the Bureau of Labor Statistics May 2019 report of wages for each occupation code. To merge the O*NET, telework, and wages measures to the death and population data, we used multiple available crosswalks from the occupational coding schemes used in each source to 2010 Census occupation codes (see Appendix “Occupation code crosswalks”).

In our presentation of the results, we focus on the occupational measures of essential sector, telework, and median annual wages, because within the three major categories of measures (sector, O*NET-based measures, and wages) these measures were the strongest predictors of COVID-19 death in the study population. Results for the remaining occupational measures are presented in Appendix Table 5.

**Appendix Table 2:** Occupation-based measures related to COVID-19 exposure risk.

| Source | Measure | Description |
| --- | --- | --- |
| California official essential worker designations<sup>1</sup> | 9 categories of essential and non-essential worker | Facilities, food/agriculture, government/community, health/emergency, manufacturing, retail, transportation/logistics, not essential, and unemployed/missing |
| Dingel and Neiman, 2020<sup>5</sup> | Telework | Classification (0/1) of feasibility of working from home for each occupation. Composite measure based on 15 O*NET job measures. |
| Bureau of Labor Statistics May 2019 National Occupational Employment and Wage Estimates | Mean, median, 10th, 25th, 75th, and 90th percentiles of annual wages; Mean, median, 10th, 25th, 75th, and 90th percentiles of hourly wages | Estimates are produced by the Occupational Employment and Wage Statistics program and provide wage estimates annually for nearly 800 occupations. Estimates are calculated with data collected from employers in all industries in urban and rural areas nationwide. |
| O*NET 2019 surveys of employees, employers, and job experts | Importance of assisting and caring for others | Work activities: Importance of “providing personal assistance, medical attention, emotional support, or other personal care to others such as coworkers, customers, or patients”. Examples of top ranking: Family medicine physicians, physical therapist assistants |
|  | Level of assisting and caring for others | Work activities: Level of “providing personal assistance, medical attention, emotional support, or other personal care to others such as coworkers, customers, or patients”. Examples of top ranking: Nurse anesthetists, sports medicine physicians |
|  | Contact with others | Work context: “How much does this job require the worker to be in contact with others (face-to-face, by telephone, or otherwise) in order to perform it?” Examples of top ranking: Allergists and immunologists, education and childcare administrators at preschools and daycares, receptionists and information clerks, telemarketers |
|  | Physical proximity | Work context: “To what extent does this job require the worker to perform job tasks in close physical proximity to other people?” Examples of top ranking: acupuncturists, choreographers, dental hygienists, sports medicine physicians |
|  | Deal with physically aggressive people | Work context: “How frequently does this job require the worker to deal with physical aggression of violent individuals?” Examples of top ranking: First-line supervisors of correctional officers, correctional officers and jailers, police and sheriff's patrol officers, psychiatric aides |
|  | Exposed to disease or infections | Work context: “How often does this job require exposure to disease/infections?” Examples of top ranking: Acute care nurses, dental hygienists, general internal medicine physicians, general pediatricians |
|  | Face-to-face discussions | Work context: “How often do you have to have face-to-face discussions with individuals or teams in this job?” Amusement and recreational attendants, buyers and purchasing agents of farm products, chief executives, electrical power-line installers and repairers, forest and conservation technicians, neurologists |
|  | Importance of interacting with computers | Work activities: Importance of “using computers and computer systems (including hardware and software) to program, write software, set up functions, enter data, or process information”. Examples of top ranking: Computer systems engineers/architects, radio frequency identification device specialists, database administrators |
|  | Level of interaction with computers | Work activities: Level of “using computers and computer systems (including hardware and software) to program, write software, set up functions, enter data, or process information”. Examples of top ranking: Computer network architects, computer systems engineers/architects, database architects |
|  | Importance of performing for or working directly with the public | Work activities: Importance of “performing for people or dealing directly with the public. This includes serving customers in restaurants and stores, and receiving clients or guests”. Examples of top ranking: Flight attendants, morticians/undertakers and funeral arrangers, demonstrators and product promoters |
|  | Level of performing for or working directly with the public | Work activities: Level of “performing for people or dealing directly with the public. This includes serving customers in restaurants and stores, and receiving clients or guests”. Examples of top ranking: Prosthodontists, judges and magistrates, morticians/undertakers and funeral arrangers |
|  | Wear specialized protective or safety equipment such as breathing apparatus, safety harness, full protection suits, or radiation protection | Work context: “How much does this job require wearing specialized protective or safety equipment such as breathing apparatus, safety harness, full protection suits, or radiation protection?” Examples of top ranking: Fiberglass laminators and fabricators, wind turbine service technicians, hazardous materials removal workers |
|  | Work with group or team | Work context: “How important is it to work with others in a group or team in this job?” Examples of top ranking: Actors, family medicine physicians, airline pilots/copilots and flight engineers |

#### Occupation code crosswalks

##### Death and American Community Survey records

Death records included open text fields for primary occupation and primary industry, described as “type of work done during most of working life”. We used the National Institute for Occupational Safety and Health’s Industry and Occupation Computerized Coding System, an automated machine-learning based system, to convert the open text fields for occupation and industry to standardized 2010 Census Codes (529 unique codes). To align the 2010 Census occupation codes in the death records with American Community Survey (ACS) which used 2018 Census occupation codes (570 unique codes), we used the Census crosswalk.^7^ When two 2018 codes were assigned to the same 2010 code, we assigned them the same 2010 code. When one 2018 code was assigned to multiple 2010, we arbitrarily selected the first, recognizing that the ultimate occupational measures used in the analysis (telework, wages, etc.) would not distinguish between these sub-categories.

##### O*NET data

To merge the O*NET data to the death/ACS data, we applied multiple crosswalks: from 2019 O*NET SOC codes (1016 unique codes) to 2018 SOC codes (867 unique codes),^8^ then from 2018 SOC codes to 2010 SOC codes (840 unique codes),^9^ then from 2010 SOC codes to 2010 Census codes (529 unique codes).^7^ At each stage, we applied the same procedures as above to assign codes that split or combined when transitioning from one coding scheme to the next.

##### Telework data

To merge the Dingel and Neiman telework data to the death/ACS data, we applied multiple crosswalks: from 2010 O*NET SOC codes (1110 unique codes) to 2010 SOC codes (840 unique codes),^10^ then from 2010 SOC codes to 2010 Census codes (529 unique codes).^7^ At each stage, we applied the same procedures to assign codes that split or combined when transitioning from one coding scheme to the next.

##### Wages data

To merge the US Bureau of Labor Statistics telework data to the death/ACS data, we applied multiple crosswalks: from the OES hybrid SOC coding scheme (824 unique codes) to 2010 SOC codes (840 unique codes),^11^ then from 2010 SOC codes to 2010 Census codes (529 unique codes).^7^ At each stage, we applied the same procedures to assign codes that split or combined when transitioning from one coding scheme to the next.

#### Supplemental results

**Appendix Table 4:** Predicted risk differences for individuals aged 18-65 per 100,000 population for the association of race/ethnicity with COVID-19 death accounting for each hypothesized mediator, by race/ethnicity and sex, California, January 1, 2020-February 12, 2021.

| Mediator | Race/<br>ethnicity | Males, Risk difference if all<br>individuals had the<br>mediator distribution of<br>Whites<br>(95%CI) | Males, % of racial/ethnic<br>differences in COVID-19<br>death eliminated if all<br>individuals had the<br>mediator distribution of<br>Whites<br>(95%CI) | Females, Risk difference if<br>all individuals had the<br>mediator distribution of<br>Whites<br>(95%CI) | Females, % of racial/<br>ethnic difference in<br>COVID-19 death<br>eliminated if all individuals<br>had the mediator<br>distribution of Whites<br>(95%CI) |
| --- | --- | --- | --- | --- | --- |
| Education | White | 0 (ref) |  | 0 (ref) |  |
|  | Latinx | -24 | 17 | -15 | 22 |
|  | Black | -10 | 11 | -4 | 8 |
|  | Asian | 6 | -- | 3 | ^ |
|  | Other | -6 | 7 | -3 | 8 |
| Essential sector | White | 0 (ref) | 0 | 0 (ref) |  |
|  | Latinx | -13 | 9 | -1 | 2 |
|  | Black | 9 | -- | 1 | -- |
|  | Asian | -1 | 3 | -1 | ^ |
|  | Other | -1 | 2 | 0 | 1 |
| Not Telework | White | 0 (ref) |  | 0 (ref) |  |
|  | Latinx | -9 | 6 | -2 | 3 |
|  | Black | 10 | -- | 1 | -- |
|  | Asian | -1 | 5 | 0 | ^ |
|  | Other | -1 | 2 | -1 | 1 |
| Annual wage | White | 0 (ref) |  | 0 (ref) |  |
|  | Latinx | -13 | 9 | -4 | 6 |
|  | Black | 2 | -- | 2 | -- |
|  | Asian | 0 | 1 | 0 | ^ |
|  | Other | -2 | 3 | -2 | 3 |
| All mediators | White | 0 (ref) |  | 0 (ref) |  |
|  | Latinx | -31 | 22 | -16 | 23 |
|  | Black | 0 | 0 | -4 | 7 |
|  | Asian | 6 | -- | 3 | ^ |
|  | Other | -3 | 4 | -4 | 9 |

Legend: --: Not applicable: mediator does not explain inequity. ^: Not applicable: no inequity at baseline. All models are adjusted for covariates age group, nativity, and region of residence. Risk differences were calculated by comparing the composition-adjusted COVID-19 mortality risk (the COVID-19 mortality risk if all racial/ethnic groups had the same distribution of covariates as Whites) to the composition- and mediator-adjusted COVID-19 mortality risk (the COVID-19 mortality risk if all racial/ethnic groups had the same distribution of covariates and education/occupation mediator(s) as Whites). We calculated the percent of the COVID-19 mortality racial/ethnic inequity that is explained by the education/occupation mediator(s) as the percentage change in predicted COVID-19 mortality when moving from the composition-adjusted scenario to the composition- and mediator-adjusted scenario.

**Appendix Table 4:** Demographic, educational, and occupational characteristics of study population by race/ethnicity.

|  | White |  | Latino |  | Black |  | Asian |  | Other |  |
| --- | --- | --- | --- | --- | --- | --- | --- | --- | --- | --- |
|  | *N* | % | *N* | % | *N* | % | *N* | % | *N* | % |
| Persons | 9,040,379 | 36% | 9,859,259 | 39% | 1,472,151 | 6% | 3,925,494 | 16% | 937,809 | 4% |
| COVID-19 death |  |  |  |  |  |  |  |  |  |  |
| No | 9,038,381 | 100% | 9,849,030 | 100% | 1,471,185 | 100% | 3,924,376 | 100% | 937,337 | 100% |
| Yes | 1,998 | 0% | 10,229 | 0% | 966 | 0% | 1,118 | 0% | 472 | 0% |
| Age (years) |  |  |  |  |  |  |  |  |  |  |
| 18-24 | 1,017,420 | 11% | 1,799,211 | 18% | 226,881 | 15% | 472,163 | 12% | 172,745 | 18% |
| 25-44 | 3,702,330 | 41% | 4,717,922 | 48% | 639,966 | 43% | 1,818,665 | 46% | 468,423 | 50% |
| 45-65 | 4,320,629 | 48% | 3,342,126 | 34% | 605,304 | 41% | 1,634,666 | 42% | 296,641 | 32% |
| Sex |  |  |  |  |  |  |  |  |  |  |
| Female | 4,407,412 | 49% | 4,852,961 | 49% | 723,166 | 49% | 2,063,142 | 53% | 468,250 | 50% |
| Male | 4,632,967 | 51% | 5,006,298 | 51% | 748,985 | 51% | 1,862,352 | 47% | 469,559 | 50% |
| Foreign-born |  |  |  |  |  |  |  |  |  |  |
| No | 8,078,201 | 89% | 5,576,110 | 57% | 1,319,700 | 90% | 1,181,970 | 30% | 773,811 | 83% |
| Yes | 962,108 | 11% | 4,283,083 | 43% | 152,429 | 10% | 2,743,509 | 70% | 163,934 | 17% |
| Missing | 70 | 0% | 66 | 0% | 22 | 0% | 15 | 0% | 64 | 0% |
| Educational attainment |  |  |  |  |  |  |  |  |  |  |
| No high school degree and no GED | 402,555 | 4% | 2,726,451 | 28% | 136,774 | 9% | 295,891 | 8% | 71,527 | 8% |
| High school degree or GED | 1,676,499 | 19% | 2,815,628 | 29% | 369,080 | 25% | 536,599 | 14% | 196,998 | 21% |
| Some college or Associate's degree | 3,006,774 | 33% | 2,993,868 | 30% | 592,886 | 40% | 946,735 | 24% | 342,069 | 36% |
| Bachelor's degree or beyond | 3,954,439 | 44% | 1,322,962 | 13% | 373,367 | 25% | 2,146,241 | 55% | 327,154 | 35% |
| Missing | 112 | 0% | 350 | 0% | 44 | 0% | 28 | 0% | 61 | 0% |
| Worker sector |  |  |  |  |  |  |  |  |  |  |
| Facilities | 733,589 | 8% | 1,508,582 | 15% | 81,139 | 6% | 169,041 | 4% | 69,032 | 7% |
| Food and agriculture | 466,679 | 5% | 1,065,046 | 11% | 63,386 | 4% | 235,104 | 6% | 58,329 | 6% |
| Government and community | 1,000,243 | 11% | 748,296 | 8% | 166,907 | 11% | 303,395 | 8% | 94,959 | 10% |
| Health or emergency | 733,815 | 8% | 602,333 | 6% | 163,842 | 11% | 440,885 | 11% | 84,654 | 9% |
| Manufacturing | 302,820 | 3% | 616,232 | 6% | 33,570 | 2% | 175,261 | 4% | 29,690 | 3% |
| Retail | 501,385 | 6% | 720,072 | 7% | 99,151 | 7% | 199,368 | 5% | 63,657 | 7% |
| Transportation and logistics | 455,806 | 5% | 957,648 | 10% | 145,460 | 10% | 177,834 | 5% | 68,379 | 7% |
| Not essential | 3,582,906 | 40% | 1,889,471 | 19% | 386,578 | 26% | 1,623,780 | 41% | 322,266 | 34% |
| Unemployed or missing | 1,263,136 | 14% | 1,751,579 | 18% | 332,118 | 23% | 600,826 | 15% | 146,843 | 16% |
| Telework-able occupation |  |  |  |  |  |  |  |  |  |  |
| Yes | 4,159,608 | 46% | 2,262,639 | 23% | 484,019 | 33% | 1,683,540 | 43% | 368,002 | 39% |
| No | 3,444,972 | 38% | 5,716,117 | 58% | 625,829 | 43% | 1,582,613 | 40% | 404,135 | 43% |
| Unemployed/not in labor force | 1,263,135 | 14% | 1,751,573 | 18% | 332,118 | 23% | 600,826 | 15% | 146,843 | 16% |
| Missing | 172,664 | 2% | 128,930 | 1% | 30,185 | 2% | 58,515 | 1% | 18,829 | 2% |
| Median annual wage for occupation |  |  |  |  |  |  |  |  |  |  |
| \$22,200 – 29,000 | 1,151,490 | 13% | 2,544,448 | 26% | 275,161 | 19% | 583,668 | 15% | 167,442 | 18% |
| \$29,001 – 39,100 | 1,136,431 | 13% | 2,065,638 | 21% | 294,545 | 20% | 477,281 | 12% | 156,869 | 17% |
| \$39,101 – 51,700 | 1,442,786 | 16% | 1,644,828 | 17% | 201,257 | 14% | 521,040 | 13% | 139,735 | 15% |
| \$51,701 – 73,800 | 1,839,359 | 20% | 1,104,127 | 11% | 203,665 | 14% | 666,910 | 17% | 155,091 | 17% |
| \$73,800 + | 2,124,201 | 23% | 712,908 | 7% | 147,163 | 10% | 1,048,661 | 27% | 164,032 | 17% |
| Unemployed/not in labor force | 1,263,135 | 14% | 1,751,573 | 18% | 332,118 | 23% | 600,826 | 15% | 146,843 | 16% |
| Missing | 82,977 | 1% | 35,737 | 0% | 18,242 | 1% | 27,108 | 1% | 7,797 | 1% |

**Appendix Table 5:** Predicted COVID-19 mortality risks for individuals aged 18-65, per 100,000 persons, by race/ethnicity and sex, if all groups had the same composition and occupational risks as Whites, California, January 1, 2020-February 12, 2021.

| Occupational measure (mediator) | Race/ethnicity | Males, Composition-adjusted | Males, Composition- and mediator-adjusted | Females, Composition-adjusted | Females, Composition- and mediator-adjusted |
| --- | --- | --- | --- | --- | --- |
| Importance of assisting and caring for others | White | 26 | 26 | 15 | 15 |
|  | Latinx | 142 | 145 | 70 | 70 |
|  | Black | 87 | 101 | 51 | 52 |
|  | Asian | 27 | 26 | 13 | 14 |
|  | Other | 84 | 87 | 46 | 46 |
| Level of assisting and caring for others | White | 26 | 26 | 15 | 15 |
|  | Latinx | 142 | 142 | 70 | 69 |
|  | Black | 87 | 100 | 51 | 52 |
|  | Asian | 27 | 26 | 13 | 14 |
|  | Other | 84 | 87 | 46 | 46 |
| Contact with others | White | 26 | 26 | 15 | 15 |
|  | Latinx | 142 | 143 | 70 | 68 |
|  | Black | 87 | 99 | 51 | 54 |
|  | Asian | 27 | 27 | 13 | 14 |
|  | Other | 84 | 87 | 46 | 46 |
| Physical proximity | White | 26 | 26 | 15 | 15 |
|  | Latinx | 142 | 143 | 70 | 69 |
|  | Black | 87 | 97 | 51 | 53 |
|  | Asian | 27 | 26 | 13 | 12 |
|  | Other | 83 | 85 | 46 | 46 |
| Deal with physically aggressive people | White | 26 | 26 | 16 | 16 |
|  | Latinx | 142 | 144 | 70 | 69 |
|  | Black | 87 | 98 | 50 | 50 |
|  | Asian | 28 | 26 | 13 | 13 |
|  | Other | 84 | 85 | 46 | 45 |
| Exposed to disease or infections | White | 26 | 26 | 16 | 16 |
|  | Latinx | 142 | 144 | 70 | 69 |
|  | Black | 88 | 101 | 51 | 52 |
|  | Asian | 30 | 28 | 13 | 14 |
|  | Other | 86 | 91 | 47 | 47 |
| Face-to-face discussions | White | 26 | 26 | 15 | 15 |
|  | Latinx | 142 | 143 | 70 | 68 |
|  | Black | 87 | 97 | 51 | 52 |
|  | Asian | 27 | 27 | 13 | 12 |
|  | Other | 84 | 87 | 46 | 46 |
| Importance of interacting with computers | White | 26 | 26 | 16 | 16 |
|  | Latinx | 142 | 132 | 70 | 67 |
|  | Black | 86 | 93 | 50 | 51 |
|  | Asian | 28 | 28 | 13 | 13 |
|  | Other | 83 | 82 | 46 | 46 |
| Level of interaction with computers | White | 26 | 26 | 15 | 15 |
|  | Latinx | 142 | 136 | 67 | 65 |
|  | Black | 85 | 93 | 48 | 49 |
|  | Asian | 26 | 26 | 12 | 12 |
|  | Other | 85 | 85 | 44 | 44 |
| Importance of performing for or working directly with the public | White | 26 | 26 | 15 | 15 |
|  | Latinx | 142 | 142 | 70 | 69 |
|  | Black | 87 | 99 | 51 | 53 |
|  | Asian | 27 | 27 | 13 | 13 |
|  | Other | 84 | 88 | 46 | 46 |
| Level of performing for or working directly with the public | White | 26 | 26 | 15 | 15 |
|  | Latinx | 140 | 140 | 70 | 70 |
|  | Black | 86 | 99 | 51 | 53 |
|  | Asian | 27 | 28 | 13 | 13 |
|  | Other | 82 | 85 | 46 | 46 |
| Wear specialized protective or safety equipment such as breathing apparatus, safety harness, full protection suits, or radiation protection | White | 28 | 28 | 16 | 16 |
|  | Latinx | 144 | 144 | 72 | 71 |
|  | Black | 92 | 109 | 51 | 52 |
|  | Asian | 28 | 29 | 14 | 13 |
|  | Other | 86 | 88 | 47 | 46 |
| Work with group or team | White | 26 | 26 | 15 | 15 |
|  | Latinx | 142 | 139 | 70 | 69 |
|  | Black | 87 | 96 | 51 | 54 |
|  | Asian | 27 | 27 | 13 | 13 |
|  | Other | 84 | 86 | 46 | 46 |
| Hourly wages: mean | White | 26 | 26 | 16 | 16 |
|  | Latinx | 143 | 131 | 71 | 67 |
|  | Black | 87 | 90 | 51 | 53 |
|  | Asian | 27 | 27 | 13 | 14 |
|  | Other | 83 | 81 | 46 | 45 |
| Hourly wages: 10<sup>th</sup> percentile | White | 26 | 26 | 16 | 16 |
|  | Latinx | 143 | 131 | 71 | 68 |
|  | Black | 87 | 89 | 51 | 54 |
|  | Asian | 27 | 28 | 13 | 14 |
|  | Other | 83 | 82 | 46 | 46 |
| Hourly wages: 25<sup>th</sup> percentile | White | 26 | 26 | 16 | 16 |
|  | Latinx | 144 | 132 | 71 | 68 |
|  | Black | 88 | 89 | 52 | 53 |
|  | Asian | 27 | 27 | 14 | 14 |
|  | Other | 84 | 82 | 47 | 46 |
| Hourly wages: Median | White | 26 | 26 | 16 | 16 |
|  | Latinx | 143 | 132 | 71 | 67 |
|  | Black | 87 | 90 | 51 | 53 |
|  | Asian | 27 | 27 | 13 | 14 |
|  | Other | 83 | 82 | 46 | 45 |
| Hourly wages: 75<sup>th</sup> percentile | White | 26 | 26 | 16 | 16 |
|  | Latinx | 143 | 131 | 71 | 67 |
|  | Black | 87 | 89 | 51 | 53 |
|  | Asian | 27 | 27 | 13 | 14 |
|  | Other | 83 | 81 | 46 | 45 |
| Hourly wages: 90<sup>th</sup> percentile | White | 26 | 26 | 16 | 16 |
|  | Latinx | 143 | 131 | 71 | 67 |
|  | Black | 87 | 91 | 51 | 53 |
|  | Asian | 27 | 27 | 13 | 14 |
|  | Other | 83 | 81 | 46 | 45 |
| Annual wages: mean | White | 26 | 26 | 15 | 15 |
|  | Latinx | 142 | 128 | 71 | 67 |
|  | Black | 87 | 88 | 51 | 52 |
|  | Asian | 27 | 27 | 14 | 14 |
|  | Other | 83 | 80 | 44 | 43 |
| Annual wages: 10<sup>th</sup> percentile | White | 26 | 26 | 15 | 15 |
|  | Latinx | 141 | 130 | 70 | 67 |
|  | Black | 87 | 89 | 51 | 53 |
|  | Asian | 28 | 28 | 14 | 14 |
|  | Other | 82 | 81 | 45 | 44 |
| Annual wages: 25<sup>th</sup> percentile | White | 26 | 26 | 15 | 15 |
|  | Latinx | 142 | 130 | 71 | 67 |
|  | Black | 87 | 89 | 52 | 52 |
|  | Asian | 28 | 28 | 14 | 14 |
|  | Other | 84 | 82 | 45 | 44 |
| Annual wages: 75<sup>th</sup> percentile | White | 26 | 26 | 15 | 15 |
|  | Latinx | 142 | 129 | 71 | 67 |
|  | Black | 87 | 88 | 51 | 52 |
|  | Asian | 27 | 27 | 14 | 14 |
|  | Other | 83 | 80 | 44 | 43 |
| Annual wages: 90<sup>th</sup> percentile | White | 26 | 26 | 15 | 15 |
|  | Latinx | 142 | 129 | 71 | 67 |
|  | Black | 87 | 90 | 51 | 52 |
|  | Asian | 27 | 27 | 14 | 14 |
|  | Other | 83 | 80 | 44 | 43 |

**Appendix Table 6:** Predicted COVID-19 mortality risks for individuals aged 18-65, per 100,000 persons, by race/ethnicity and sex, if all groups had the same composition and educational/occupational risks as Whites, sensitivity analysis results based on logistic regression, California, January 1, 2020-February 12, 2021.

| Sex, race/<br>ethnicity | Composition-adjusted<br>COVID-19 mortality risk | Composition- and<br>education-adjusted<br>COVID-19 mortality<br>risk<br>(% change relative to<br>composition-adjusted) | Composition- and<br>worker sector-adjusted<br>COVID-19 mortality<br>risk<br>(% change relative to<br>composition-adjusted) | Composition- and<br>telework-adjusted<br>COVID-19 mortality<br>risk<br>(% change relative to<br>composition-adjusted) | Composition- and<br>wages-adjusted COVID-19 mortality risk<br>(% change relative to<br>composition-adjusted) |
| --- | --- | --- | --- | --- | --- |
| Female |  |  |  |  |  |
| White | 15 | 15 | 15 | 15 | 15 |
| Latinx | 63 | 47 | 63 | 61 | 61 |
| Black | 51 | 49 | 52 | 53 | 61 |
| Asian | 18 | 20 | 17 | 16 | 18 |
| Other | 45 | 41 | 46 | 47 | 45 |
| Male |  |  |  |  |  |
| White | 26 | 26 | 26 | 26 | 26 |
| Latinx | 122 | 100 | 111 | 115 | 111 |
| Black | 87 | 77 | 103 | 101 | 91 |
| Asian | 48 | 55 | 49 | 48 | 49 |
| Other | 85 | 80 | 86 | 84 | 83 |

Legend: CI: confidence interval. Composition-adjusted rates indicate the predicted COVID-19 mortality risk if all racial/ethnic groups had the same distribution of age, nativity, and region of residence as Whites.

## References

1. Centers for Disease Control and Prevention. Risk for COVID-19 Infection, Hospitalization, and Death By Race/Ethnicity [Internet]. Centers for Disease Control and Prevention. 2020 [cited 2021 Aug 2]. Available from: https://www.cdc.gov/coronavirus/2019-ncov/covid-data/investigations-discovery/hospitalization-death-by-race-ethnicity.html

2. Tai DBG, Shah A, Doubeni CA, Sia IG, Wieland ML. The Disproportionate Impact of COVID-19 on Racial and Ethnic Minorities in the United States. Clinical Infectious Diseases. 2021 Feb 15;72(4):703–6.

3. Mackey K, Ayers CK, Kondo KK, Saha S, Advani SM, Young S, et al. Racial and Ethnic Disparities in COVID-19–Related Infections, Hospitalizations, and Deaths. Ann Intern Med. 2020 Dec 1;174(3):362–73.

4. Azar KMJ, Shen Z, Romanelli RJ, Lockhart SH, Smits K, Robinson S, et al. Disparities In Outcomes Among COVID-19 Patients In A Large Health Care System In California. Health Affairs. 2020 May 21;39(7):1253–62.

5. Escobar GJ, Adams AS, Liu VX, Soltesz L, Chen Y-FI, Parodi SM, et al. Racial Disparities in COVID-19 Testing and Outcomes. Ann Intern Med [Internet]. 2021 Feb 9 [cited 2021 Feb 8]; Available from: https://www.acpjournals.org/doi/10.7326/M20-6979

6. Muñoz-Price LS, Nattinger AB, Rivera F, Hanson R, Gmehlin CG, Perez A, et al. Racial Disparities in Incidence and Outcomes Among Patients With COVID-19. JAMA Network Open. 2020 Sep 25;3(9):e2021892–e2021892.

7. McClure ES, Vasudevan P, Bailey Z, Patel S, Robinson WR. Racial Capitalism Within Public Health—How Occupational Settings Drive COVID-19 Disparities. American Journal of Epidemiology. 2020 Nov 2;189(11):1244–53.

8. Laster Pirtle WN. Racial Capitalism: A Fundamental Cause of Novel Coronavirus (COVID-19) Pandemic Inequities in the United States. Health Educ Behav. 2020 Aug 1;47(4):504–8.

9. Bhala N, Curry G, Martineau AR, Agyemang C, Bhopal R. Sharpening the global focus on ethnicity and race in the time of COVID-19. The Lancet. 2020 May 30;395(10238):1673–6.

10. Hawkins D. Differential occupational risk for COVID-19 and other infection exposure according to race and ethnicity. American Journal of Industrial Medicine. 2020;63(9):817–20.

11. Chen Y-H, Glymour M, Riley A, Balmes J, Duchowny K, Harrison R, et al. Excess mortality associated with the COVID-19 pandemic among Californians 18–65 years of age, by occupational sector and occupation: March through November 2020. PLOS ONE. 2021 Jun 4;16(6):e0252454.

12. Selden TM, Berdahl TA. COVID-19 And Racial/Ethnic Disparities In Health Risk, Employment, And Household Composition. Health Affairs. 2020 Jul 14;39(9):1624–32.

13. Benitez J, Courtemanche C, Yelowitz A. Racial and Ethnic Disparities in COVID-19: Evidence from Six Large Cities. J Econ Race Policy. 2020 Dec 1;3(4):243–61.

14. Figueroa JF, Wadhera RK, Lee D, Yeh RW, Sommers BD. Community-Level Factors Associated With Racial And Ethnic Disparities In COVID-19 Rates In Massachusetts. Health Affairs. 2020 Nov 1;39(11):1984–92.

15. Figueroa JF, Wadhera RK, Mehtsun WT, Riley K, Phelan J, Jha AK. Association of race, ethnicity, and community-level factors with COVID-19 cases and deaths across U.S. counties. Healthcare. 2021 Mar 1;9(1):100495.

16. Reitsma MB, Claypool AL, Vargo J, Shete PB, McCorvie R, Wheeler WH, et al. Racial/Ethnic Disparities In COVID-19 Exposure Risk, Testing, And Cases At The Subcounty Level In California. Health Affairs. 2021 Jun 1;40(6):870–8.

17. Bui DP. Racial and Ethnic Disparities Among COVID-19 Cases in Workplace Outbreaks by Industry Sector — Utah, March 6–June 5, 2020. MMWR Morb Mortal Wkly Rep [Internet]. 2020 [cited 2021 Jun 28];69. Available from: https://www.cdc.gov/mmwr/volumes/69/wr/mm6933e3.htm

18. Chen Y-H, Glymour MM, Catalano R, Fernandez A, Nguyen T, Kushel M, et al. Excess Mortality in California During the Coronavirus Disease 2019 Pandemic, March to August 2020. JAMA Internal Medicine. 2021 May 1;181(5):705–7.

19. Laster Pirtle WN, Wright T. Structural Gendered Racism Revealed in Pandemic Times: Intersectional Approaches to Understanding Race and Gender Health Inequities in COVID-19. Gender & Society. 2021 Apr 1;35(2):168–79.

20. Hawkes S, Buse K. COVID-19 and the gendered markets of people and products: explaining inequalities in infections and deaths. Canadian Journal of Development Studies / Revue canadienne d’études du développement. 2020 Oct 23;0(0):1–18.

21. Jones CP. Invited Commentary: “Race,” Racism, and the Practice of Epidemiology. American Journal of Epidemiology. 2001 Aug 15;154(4):299–304.

22. State of California. Essential workforce [Internet]. 2020 [cited 2021 Jul 6]. Available from: https://covid19.ca.gov/essential-workforce/

23. Naimi AI, Schnitzer ME, Moodie EEM, Bodnar LM. Mediation Analysis for Health Disparities Research. American Journal of Epidemiology. 2016 Aug 15;184(4):315–24.

24. Snowden JM, Rose S, Mortimer KM. Implementation of G-Computation on a Simulated Data Set: Demonstration of a Causal Inference Technique. Am J Epidemiol. 2011 Apr 1;173(7):731–8.

25. Angrist J, Pischke J-S. 3.1 Regression Fundamentals. In: Mostly Harmless Econometrics. Princeton, New Jersey: Princeton University Press; 2008.

26. Hawkins RB, Charles EJ, Mehaffey JH. Socio-economic status and COVID-19–related cases and fatalities. Public Health. 2020 Dec 1;189:129–34.

27. Chadeau-Hyam M, Bodinier B, Elliott J, Whitaker MD, Tzoulaki I, Vermeulen R, et al. Risk factors for positive and negative COVID-19 tests: a cautious and in-depth analysis of UK biobank data. International Journal of Epidemiology. 2020 Oct 1;49(5):1454–67.

28. Abedi V, Olulana O, Avula V, Chaudhary D, Khan A, Shahjouei S, et al. Racial, Economic, and Health Inequality and COVID-19 Infection in the United States. J Racial and Ethnic Health Disparities. 2021 Jun 1;8(3):732–42.

29. Bailey ZD, Krieger N, Agénor M, Graves J, Linos N, Bassett MT. Structural racism and health inequities in the USA: evidence and interventions. The Lancet. 2017 Apr 8;389(10077):1453–63.

30. Alobuia WM, Dalva-Baird NP, Forrester JD, Bendavid E, Bhattacharya J, Kebebew E. Racial disparities in knowledge, attitudes and practices related to COVID-19 in the USA. Journal of Public Health. 2020 Aug 18;42(3):470–8.

31. Zuvekas SH, Taliaferro GS. Pathways To Access: Health Insurance, The Health Care Delivery System, And Racial/Ethnic Disparities, 1996–1999. Health Affairs. 2003 Mar 1;22(2):139–53.

32. Chen J, Vargas-Bustamante A, Mortensen K, Ortega AN. Racial and Ethnic Disparities in Health Care Access and Utilization Under the Affordable Care Act. Med Care. 2016 Feb;54(2):140–6.

33. Patler C, Gleeson S, Schonlau M. Contesting Inequality: The Impact of Immigrant Legal Status and Education on Legal Knowledge and Claims-Making in Low-Wage Labor Markets. Social Problems [Internet]. 2020 Oct 14 [cited 2021 Aug 18];(spaa029). Available from: https://doi.org/10.1093/socpro/spaa029

34. Pham O, Jun 30 NPP, 2021. Latest Data on COVID-19 Vaccinations by Race/Ethnicity [Internet]. KFF. 2021 [cited 2021 Jul 7]. Available from: https://www.kff.org/coronavirus-covid-19/issue-brief/latest-data-on-covid-19-vaccinations-race-ethnicity/

35. Riley AR, Chen Y-H, Matthay EC, Glymour MM, Torres JM, Fernandez A, et al. Excess death among Latino people in California during the COVID-19 pandemic. medRxiv. 2021 Jan 25;2020.12.18.20248434.

36. Wrigley-Field E, Garcia S, Leider JP, Robertson C, Wurtz R. Racial Disparities in COVID-19 and Excess Mortality in Minnesota. Socius. 2020 Jan 1;6:2378023120980918.

37. Braveman PA, Cubbin C, Egerter S, Chideya S, Marchi KS, Metzler M, et al. Socioeconomic Status in Health ResearchOne Size Does Not Fit All. JAMA. 2005 Dec 14;294(22):2879–88.

## References

1 Chen Y-H, Glymour M, Riley A, et al. Excess mortality associated with the COVID-19 pandemic among Californians 18–65 years of age, by occupational sector and occupation: March through November 2020. PLOS ONE 2021; 16: e0252454.

2 State of California. Essential workforce. 2020. https://covid19.ca.gov/essential-workforce/ (accessed July 6, 2021).

3 Hawkins D. Differential occupational risk for COVID-19 and other infection exposure according to race and ethnicity. American Journal of Industrial Medicine 2020; 63: 817–20.

4 Baker MG. Nonrelocatable Occupations at Increased Risk During Pandemics: United States, 2018. Am J Public Health 2020; 110: 1126–32.

5 Dingel JI, Neiman B. How many jobs can be done at home? Journal of Public Economics 2020; 189: 104235.

6 Baker MG. Who cannot work from home? Characterizing occupations facing increased risk during the COVID-19 pandemic using 2018 BLS data. medRxiv 2020; : 2020.03.21.20031336.

7 US Census Bureau. 2018 Census Occupation Code List with Crosswalk. 2020; published online Aug 31. https://www2.census.gov/programs-surveys/demo/guidance/industry-occupation/2018-occupation-code-list-and-crosswalk.xlsx (accessed July 21, 2021).

8 O*NET Resource Center. Crosswalk O*NET-SOC 2019 to 2018 SOC - Occupational Listings at O*NET Resource Center. https://www.onetcenter.org/taxonomy/2019/soc.html (accessed July 21, 2021).

9 US Bureau of Labor Statistics. Crosswalk from the 2010 SOC to the 2018 SOC. 2017; published online Dec 6. https://www.bls.gov/soc/2018/soc_2010_to_2018_crosswalk.xlsx (accessed July 21, 2021).

10 O*NET Resource Center. Updating the O*NET-SOC Taxonomy: Incorporating the 2010 SOC Structure at O*NET Resource Center. 2010; published online Dec. https://www.onetcenter.org/reports/Taxonomy2010.html (accessed July 21, 2021).

11 US Bureau of Labor Statistics. OES 2019 Hybrid Structure. 2020; published online March 31. https://www.bls.gov/oes/oes_2019_hybrid_structure.xlsx.

